# Neurological, neuropsychiatric and psychiatric symptoms during COVID-19 infection and after recovery: a systematic review of observational studies

**DOI:** 10.1101/2021.07.02.21259902

**Authors:** Maria Stavrou, Solomis Solomou, Anthousa Kythreotou, Antreas Ioannou, Eva Lioutas, Joanna Lioutas, Peter Karayiannis

**Affiliations:** Centre for Clinical Brain Sciences, University of Edinburgh, United Kingdom; Royal Free Hospital, London, United Kingdom; Internal Medicine Department, Nicosia General Hospital, Nicosia, Cyprus; Microbiology/Molecular Virology Department, University of Nicosia Medical School, Nicosia, Cyprus

**Author notes:** Corresponding author: Dr Maria Stavrou.

## Abstract

**Background:** The SARS-CoV-2 virus causes a wide spectrum of disease severity. Initial manifestations include fever, dry cough, and constitutional symptoms, which may progress to respiratory disease. There may also be neurological and psychiatric manifestations, involving both the central and peripheral nervous system.

**Methods:** We performed a literature search of the databases PubMed, EMBASE, The Cochrane Library and Web of Science for observational studies reporting neurological, psychiatric, and neuropsychiatric effects of COVID-19. This was followed by a narrative synthesis to summarise the data and discuss neuropsychiatric associations, symptom severity, management, and recovery.

**Findings:** The most frequently reported neurological symptoms were ageusia, hyposmia/anosmia, dizziness, headache, and loss of consciousness. Statistically significant relationships were noted between Asian ethnicity and peripheral neuropathy (p=0.0001) and neuro-syndromic symptoms (p=0.001). ITU admission was found to have a statistically significant relationship with male sex (p=0.024). Depression and anxiety were also identified both during and after infection. The most frequent treatments used were intravenous immunoglobulins, followed by antibiotics, antivirals, and hydroxychloroquine; with mean treatment duration of 6 days.

**Interpretation:** Various neuropsychiatric symptoms have been associated with COVID-19 infection. More studies are required to further our knowledge in the management of neurological and psychiatric symptoms during and after COVID-19 infection

## INTRODUCTION

Severe acute respiratory syndrome coronavirus 2 (SARS-CoV-2) is a novel virus, initially discovered in the city of Wuhan, China.^1^ SARS-CoV-2 causes coronavirus disease (COVID-19), which has led to an ongoing global pandemic. Despite belonging to the coronavirus family, which usually cause self-limiting upper respiratory tract infections, SARS-CoV-2 is often more virulent than most coronaviruses and may lead to severe respiratory disease.^2^

The mechanism of action for SARS-CoV-2 may relate to a specific tropism for respiratory tract mucosal cells through the attachment of viral surface proteins to angiotensin-converting enzyme (ACE) 2 receptors.^3^ After infection, the virus causes a wide spectrum of disease severity, with most patients suffering a mild self-limiting disease. Initial manifestations include fever, dry cough and constitutional symptoms (headache, fatigue, myalgia, arthralgia), progressing to respiratory disease of mild to moderate severity.^2,4^ Other disease manifestations include gastrointestinal symptoms (nausea, vomiting, diarrhoea), sore throat, skin rashes, anosmia, ageusia, and chest pain.^5^ In patients with underlying comorbidities or advanced age, the infection may be complicated with acute respiratory distress syndrome (ARDS), acute renal failure, sepsis, multi-organ failure and death.^6,7^

As the pandemic of COVID-19 persists, the knowledge of the clinical disease spectrum is still unfolding. Medical literature of COVID-19 infected patients reveals a variety of extra-pulmonary organ involvement.^8^ Among these, COVID-19 has been associated with several neurological and psychiatric effects, involving both the central and peripheral nervous system.^9^

## METHODS

This systematic review follows the Preferred Reporting Items for Systematic reviews and MetaAnalyses (PRISMA) statement^10^ and was registered in the PROSPERO International Prospective Register of Systematic Reviews (number CRD42020203770 at www.crd.york.ac.uk/PROSPERO).

### Search strategy

The literature search was performed in August 2020 using the databases PubMed, EMBASE, The Cochrane Library and Web of Science, from their respective inception dates. The following search terms were used:

(Neuro* OR Nervous OR Psychiatry* OR Mental) AND (COVID OR Corona*)

The search strategies incorporated both medical subject headings (MeSH) and free-text terms, which were adapted according to the database searched. Grey literature was also searched. Reference lists of the identified papers and reviews were hand-searched. Publication languages included English and Greek. There were no publication period restrictions.

### Inclusion and exclusion criteria

Included studies were observational studies reporting neurological, psychiatric, and neuropsychiatric effects of COVID-19. The included participants were COVID-19 patients of any ethnic origin, sex/gender, age, country, and were either actively infected from COVID-19 at the time of the study or had recovered. We did not include studies examining psychiatric effects on the general population as an indirect result of the pandemic.

### Main outcomes

The main outcomes included neurological, psychiatric, and neuropsychiatric effects of COVID-19, either based on clinical diagnosis or relevant diagnostic questionnaires. Information about recovery and treatment was reported when available.

### Screening

Titles were screened for inclusion, followed by screening of abstracts, and then content. One author (SS) screened the papers, and any disagreements were resolved by discussion with the review’s primary author (MS) and the other authors.

### Data extraction

The Cochrane good practice data extraction form was used for data extraction. Data extraction from reviews involved the NICE extraction form, and the data were extracted in an electronic format.

### Risk of bias/quality assessment

The quality and risk of bias were assessed by the Mixed Methods tool for Appraisal (MMAT). The guidance from the Centre for Reviews and Dissemination was used for the appraisal of review papers. Discrepancies were resolved by discussion within the authors’ team.

### Strategy for data synthesis

We performed a narrative synthesis review of original studies and reviews reporting neurological, psychiatric, and neuropsychiatric effects in COVID-19 patients. We summarised the data and discussed: (1) neuropsychiatric associations, (2) symptom severity, (3) management, and (4) recovery. Information from the various identified studies was analysed, summarised, and compared.

## RESULTS

Following our literature search, we identified a total of 7,460 papers. After removing the duplicated and irrelevant papers, 328 full text articles remained to be assessed for eligibility using the inclusion and exclusion criteria. Of these, 313 studies were included in the final narrative synthesis: specifically, 307 studies for neurological symptoms and 7 studies for psychiatric symptoms, as shown in Figure 1. A total of 15 full text papers were excluded as they were either not relevant (n=4) or unrelated to COVID-19 infection (n=11).

**Figure 1.**
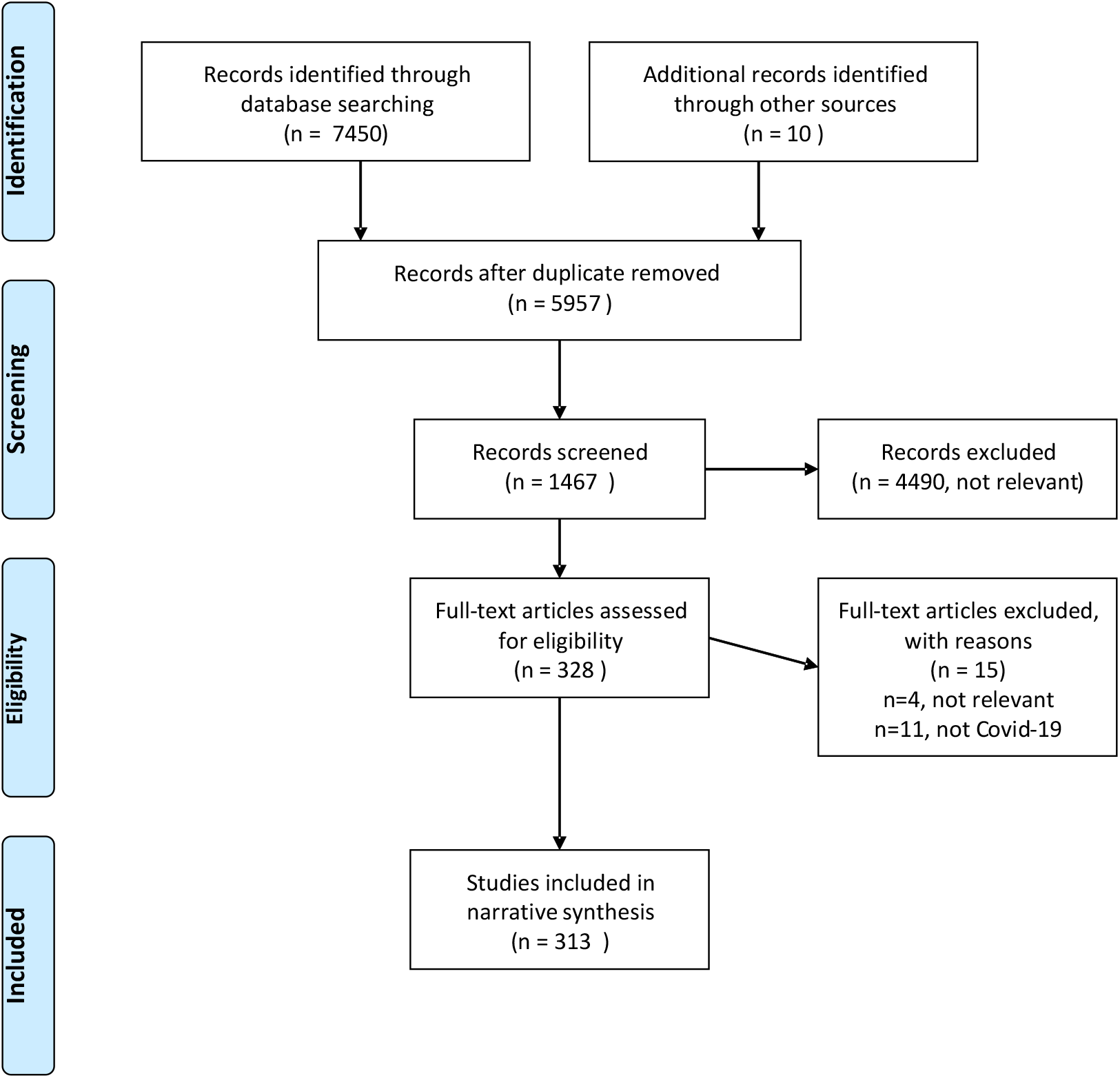
PRISMA flowchart of selected studies.

### Neurological symptoms

#### Data synthesis

A total of 307 studies for neurological symptoms were included in the narrative synthesis, as mentioned above, of which 202 were case reports, 53 case series, 2 retrospective studies, 21 cohort studies, 15 systematic reviews, 8 cross-sectional studies, 3 case-control studies, and 3 retrospective case series. A summary of the studies included in the systematic review is shown in Table 1, and a complete list of the studies is provided in Supplementary Material 1. The mean age of the patients included was 55.11 (±17.91) years. Most of the patients in our cohort were males (61%) and the majority of the participants were Asians (57%).

**Table 1.** Summary of studies included in the systematic review for neurological symptoms.

|  | Number of studies | Total sample size | Average sample size | Average female % | Average male % |
| --- | --- | --- | --- | --- | --- |
| **Type of Study** |  |  |  |  |  |
| Case report | 202 | 2,104 | 18 | 38 | 62 |
| Cross-sectional | 8 | 10,623 | 1,328 | 48 | 52 |
| Retrospective study | 2 | 223 | 112 | 46 | 54 |
| Case series | 53 | 3,447 | 75 | 27 | 73 |
| Retrospective case series | 3 | 578 | 193 | 40 | 60 |
| Systematic review | 15 | 36,724 | 3,023 | 41 | 59 |
| Cohort study | 21 | 6,180 | 294 | 34 | 66 |
| Case control | 3 | 218 | 73 | 37 | 63 |
| **Total of all studies** | **307** | **60,097** | **640** | **39** | **61** |

#### Clinical Presentation

A total of 107 studies (42.7%), involving 26,758 patients, included a full account of neurological symptoms experienced by the participants following COVID-19 infection. Table 2 presents the frequency of symptoms and their resolution. The most reported symptoms were ageusia (n=390), hyposmia/anosmia (n=480), dizziness (n=230), headache (n=860), and loss of consciousness (n=310).

**Table 2.** Frequency and recovery rates of different COVID-19 neurological presentations

|  | **n patients** | **n patients resolved** | **% of patients whose symptoms resolved** |
| --- | --- | --- | --- |
| **Clinical Presentation** |  |  |  |
| **Headache** | 860 | 690 | 80% |
| **Skeletal symptoms** | 730 | 700 | 100% |
| **Acute CVA** | 500 | 370 | 74% |
| **Hyposmia/anosmia** | 480 | 420 | 87% |
| **Ageusia** | 390 | 360 | 92% |
| **Meningitis-encephalitis** | 380 | 330 | 86% |
| **Encephalopathies** | 380 | 300 | 78% |
| **Neurological syndromes** | 320 | 230 | 71% |
| **Loss of consciousness** | 310 | 290 | 93% |
| **Dyspnoea** | 280 | .. | .. |
| **Seizure** | 260 | 230 | 88% |
| **Dizziness** | 230 | 200 | 86% |
| **Fatigue** | 130 | .. | .. |
| **Fever** | 110 | .. | .. |
| **Peripheral nervous system** | 90 | 60 | 66% |
| **Dry cough** | 81 | .. | .. |
| **Cerebellar symptoms** | 70 | 60 | 85% |
| **Spinal cord syndromes** | 30 | 10 | 33% |
| **Flu-Like symptoms** | 30 | .. | .. |
| **Dysphagia/odynophagia** | 30 | .. | .. |
| **Anorexia** | 30 | .. | .. |
| **Abdominal pain** | 20 | .. | .. |
| **GI symptoms** | 18 | .. | .. |
| **Haemoptysis** | 10 | .. | .. |

Moreover, a significant number of patients experienced severe neurological manifestations, such as seizures (n=260), acute cerebrovascular events (n=500), cerebellar syndromes (n=70), peripheral neuropathies (n=90), meningitis/encephalitis (n=380), encephalopathies (n=380), neurological syndromes such as Guillain-Barre syndrome (n=320), and spinal cord syndromes (n=30).

A statistically significant relationship was noted between ethnicity and peripheral neuropathy (p=0.0001) as well as between ethnicity and neuro-syndromic symptoms (p=0.001), with Asian patients being more likely to experience these symptoms. Both sexes were statistically as likely to present with symptoms of ageusia (p=0.0001), dizziness (p=0.033), gastrointestinal symptoms (p=0.0001), and anorexia (p=0.0001). However, flu-like symptoms were statistically more prevalent in females (p=0.008), whereas hyposmia (p=0.037) and haemoptysis (p=0.0001) was more frequent in males.

Following recovery from COVID-19 infection, a large proportion of patients demonstrated a complete resolution of their symptoms. Specifically, patients presenting with loss of consciousness and ageusia reported the highest resolution rates (93% and 92% respectively), while the patients that experienced spinal cord syndromes had the lowest resolution rates of their symptoms (33%).

#### Treatments

The most frequent treatments used in the studies analysed were intravenous immunoglobulins (IVIG) (20.17%), followed by antibiotics such as azithromycin (19.29%), antivirals (14.91%), and hydroxychloroquine (10.52%). However, a combination of therapies was required for treatment in some patients. Figure 2 illustrates the different types of drugs that the COVID-19 patients received during their admission and how the drug therapy is markedly heterogeneous among this group of patients.

**Figure 2.**
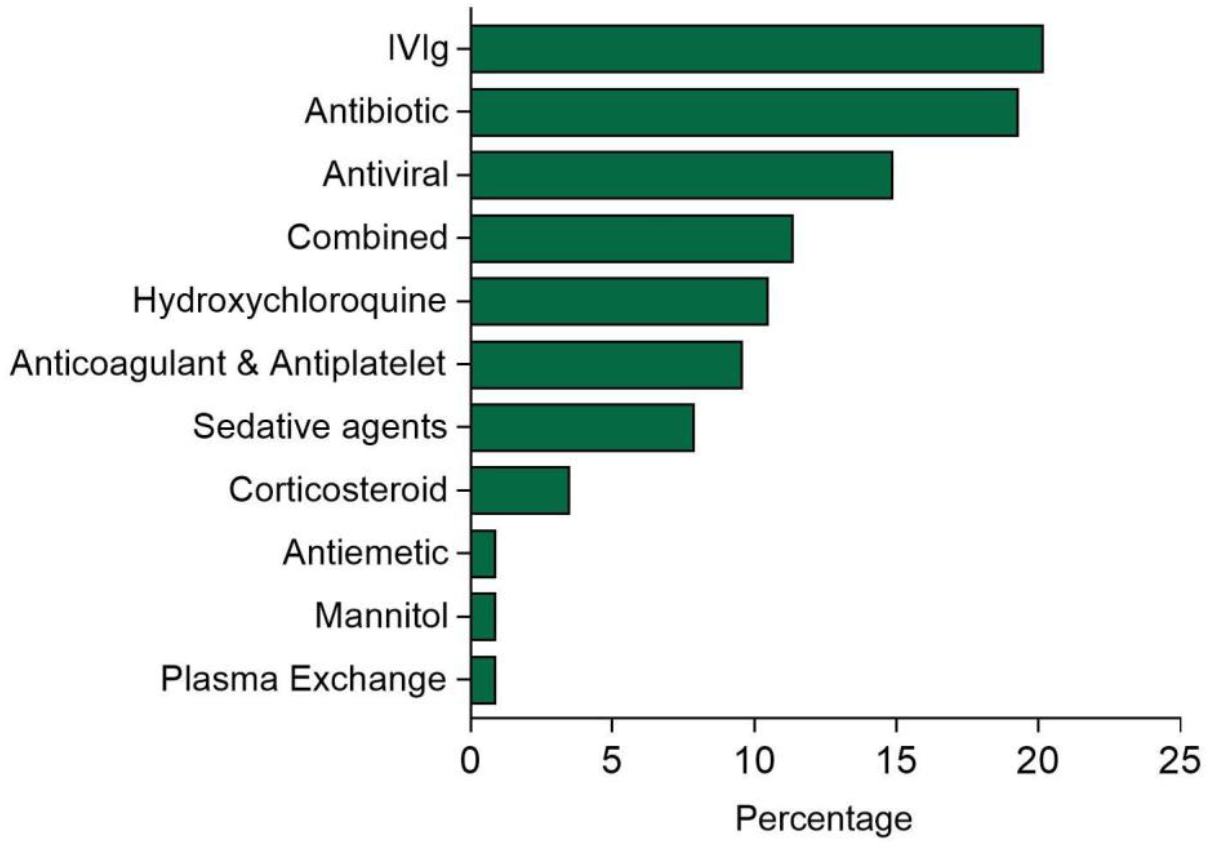
Drug type administered to COVID-19 patients.

The most common route of drug administration was intravenous (65%), although oral drug administration and intramuscular injections were also utilised. Patients received treatment for a mean duration of 6 (±4) days.

#### Prognosis

Patients admitted to an Intensive Therapy Unit (ITU) were reported in 126 studies. Figure 3 shows the different types of management that patients received when admitted to ITU and illustrates that the most common cause of ITU admission was the need for respiratory support with intubation and mechanical ventilation (84% of the cases).

**Figure 3.**
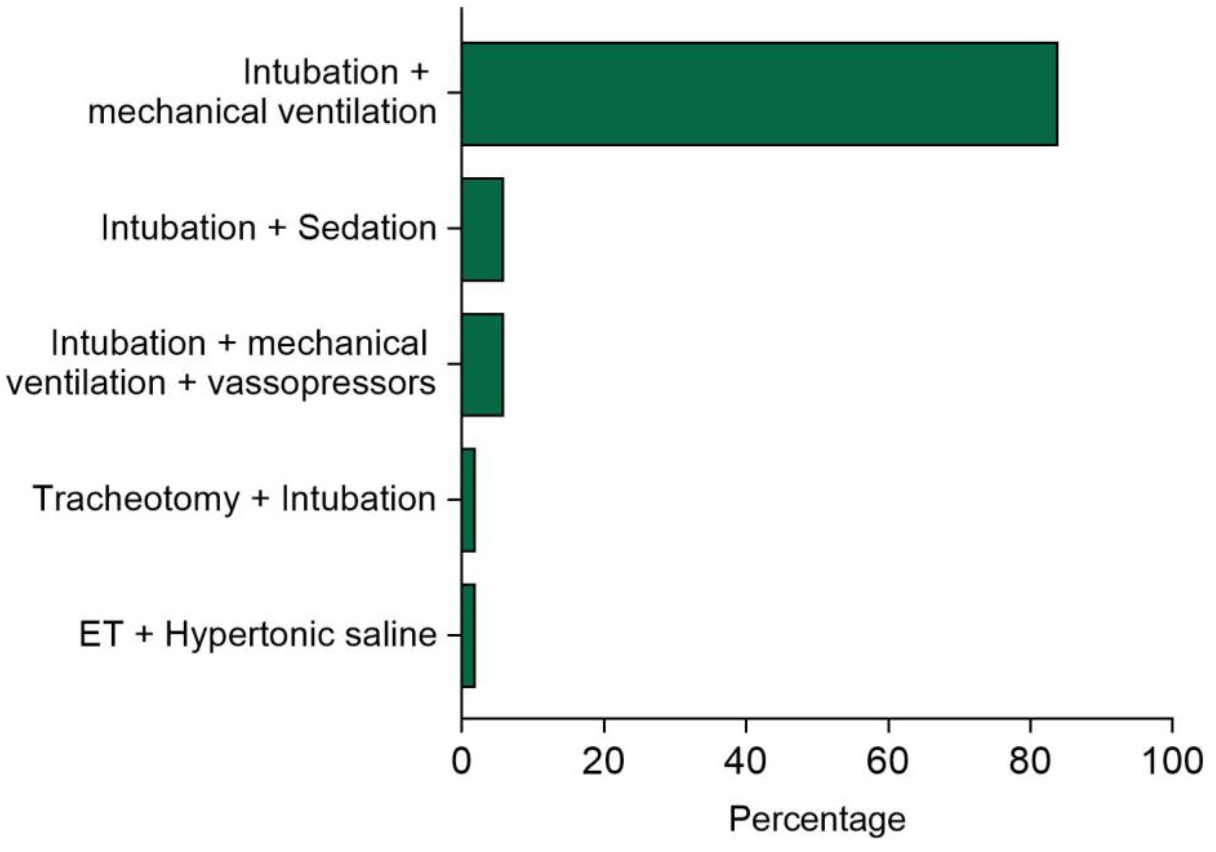
Types of ITU management received by patients.

ITU admission was found to have a statistically significant relationship with males (p=0.024), but not age. Interestingly, there was a statistically significant relationship with ITU admission and symptoms of hyposmia/anosmia (p=0.0001), headache (p=0.035), acute CVA (p=0.0001), seizure (p=0.001), meningitis (p=0.034), and encephalopathies (p=0.0001).

### Psychiatric symptoms

We identified seven studies reporting psychiatric effects, of which five were cross-sectional studies, one was a retrospective cohort study, and one was a case report. Details of the six studies are reported in Table 3. The studies involved 299,000 patients in total, of which 44% were male and 56% were female. Half of the studies were reported in China. Three studies involved 171 patients in hospital settings while having active COVID-19 infection, three studies involved 498 patients at home after recovery, and one study involved 62,354 patients covering both inpatients during infection and those at home after recovery. All studies identified depression and anxiety as being relevant to COVID-19 infection, both during and after infection. Additionally, one study reported suicidality during infection, two studies reported post-traumatic stress disorder after infection, one study suggested obsessive-compulsive disorder after infection, one study suggested insomnia after infection, one study suggested a higher incidence of psychosis, and two studies suggested a higher incidence of dementia diagnosis as being relevant to having been diagnosed with COVID-19.

**Table 3.** Studies reporting psychiatric effects related to COVID-19 infection.

| Study | Type | Country | Setting | Number, age | Condition/scale | Results | Conclusions |
| --- | --- | --- | --- | --- | --- | --- | --- |
| **Epstein et al.<sup>11</sup>** | Case report | Israel | Isolation, hospital ward | N=1; Males: n=1<br>Age=34 | Anxiety, insomnia | Suicide attempted on day 7 | Regular screening needed for depression, anxiety and suicidality |
| **Mazza et al.<sup>12</sup>** | Cross-sectional | Italy | Home (post hospital discharge) | N=402; Males: n=265, Females: n=137; Mean age=58 | IES-R, PCL-5, ZZSDS, OCI, BDI-13, STAI-Y, MOS-SS, WHIIRS | A significant proportion of patients self-rated in the psycho-pathological range: 28% for PTSD, 31% for depression, 42% for anxiety, 20% for OC symptoms, and 40% for insomnia. | Regular assessment of psychopathology of COVID-19 survivor needed to diagnose and treat emergent psychiatric conditions |
| **Yuan et al.<sup>13</sup>** | Cross-sectional | China | Home (post hospital discharge) | N=96; Males: n=47, Females: n=49; 18-45 years: n=44, >46 years: n=52 | PTSD-SS, ZSDS, ZSAS | 44% reported depressive symptoms. No significant correlation of depression with sex, age, comorbidity, severity of initial infection, and duration of initial illness. There were changes in immune system function. | Appropriate psychological interventions are necessary for COVID-19 survivors |
| **Taquet et al.<sup>14</sup>** | Cross-sectional | USA | Various, during the first 14 to 90 days after diagnosis | N=62,354; Males: n=27,525, Females: n=34,564, Other: n=265; Mean age=49.3 | Incidence of psychiatric diagnosis, including dementia | Incidence of any psychiatric diagnosis= 18.1%; Incidence of a first diagnosis of dementia= 1.6% | There is psychiatric morbidity to be anticipated in survivors of COVID-19 and for which services need to plan. |
| **Kong et al.<sup>15</sup>** | Cross-sectional | China | Inpatient ward | N=144; Males: n=70, Females: n=74; Mean age= 50 | HADS, PSSS | 34.72% and 28.47% had symptoms of anxiety or depression, respectively. Less social support was correlated with more anxious ($r=-0.196$, $p<0.05$) and depressive ($r=-0.360$, $p<0.05$) symptoms | Mental health assessments and appropriate interventions are essential part of inpatient clinical care for those at risk. |
| **Yang et al.<sup>16</sup>** | Cross-sectional | China | Inpatient, isolation ward | N=26 | HAMD, HAMA | Depression and anxiety scores were higher in the COVID-19 group compared to other groups (health and general pneumonia). Scores decreased with psychological intervention one week later. | Mental health interventions for inpatients can be beneficial for stress and anxiety during COVID-19 infection. |
| **Taquet et al.<sup>17</sup>** | Retrospective cohort study | USA | 6-month outpatient follow-up | N=236 379<br>Age>10 years | Incidence of psychiatric presentations obtained from the TriNetX electronic health records network | The whole COVID-19 cohort had estimated incidences of 0.67% (0.59–0.75) for dementia, 17.39% (17.04–17.74) for anxiety disorder, and 1.40% (1.30–1.51) for psychotic disorder. In the group with ITU admission, estimated incidences were 1.74% (1.31–2.30) for dementia, 19.15% (17.90–20.48) for anxiety disorder, and 2.77% (2.31–3.33) for psychotic disorder. | There is substantial psychiatric morbidity in the 6 months after COVID-19 infection. Risks were greatest in patients who had severe COVID-19. These results must be taken into account for in service planning and identification of research priorities. |

Note: Impact of Events Scale-Revised (IES-R), PTSD Checklist for DSM-5 (PCL-5), Zung Self-Rating Depression Scale (ZSDS), 13-item Beck's Depression Inventory (BDI-13), State-Trait Anxiety Inventory form Y (STAI-Y), Medical Outcomes Study Sleep Scale (MOS-SS), Women's Health Initiative Insomnia Rating Scale (WHIIRS), and Obsessive-Compulsive Inventory (OCI), post-traumatic stress disorder self-rating scale (PTSD-SS), Zung anxiety self-rating scale (ZSAS), Hospital Anxiety and Depression Scale (HADS), Perceived Social Support Scale (PSSS), Hamilton depression scale (HAMD), and Hamilton anxiety scale (HAMA).

## DISCUSSION

The literature published on the neurological symptoms observed in patients with COVID-19 is vast. Through our review, we aimed to summarise all available literature, as well as include more recent studies that older reviews may not have included. Our review specifically served to identify and examine the frequency and severity of these symptoms through combining this existing literature. In total, 307 neurological studies covering 60,097 patients, were included in this systematic review, which has shown that COVID-19 is associated with a large variety of neurological symptoms. The most frequently reported symptoms included ageusia, hyposmia/anosmia, dizziness, headache, and loss of consciousness. These symptoms are not specific to SARS-CoV-2 infection and are of low severity, however they may suggest neurotropism. They also associate with high resolution rates (all>80%). The most common severe neurological complication of COVID-19 was acute cerebrovascular events. This result is in keeping with other systematic reviews.^18,19^

Direct neurological damage including ischemic strokes, meningitis/encephalitis, or Guillain-Barre syndrome are relatively common extra-pulmonary neurological presentations according to our review. These results should be the springboard for further research efforts aiming to distinguish whether these neurological entities are a consequence of direct brain injury/infection or an interaction with other vascular comorbidities of patients suffering severe/critical COVID-19 disease.

A significant proportion of COVID-19 patients were asymptomatic due to the course of SARS-CoV-2 infection. In addition, patients may not present with respiratory symptoms or fever but still have initial neurological manifestations. Thus, when patients present with neurological symptoms, despite the absence of respiratory symptoms, clinicians should maintain a high level of clinical suspicion for the possibility of underlying COVID-19 asymptomatic infection.

The resolution rates of neurological symptoms also varied. Patients presenting with loss of consciousness and ageusia reported the highest resolution rates (93% and 92% respectively), with ageusia resolution rates being 100% in one study.^20^ On the other hand, patients who experienced spinal cord syndromes, such as acute myelitis, had the lowest resolution rates of their symptoms (33%). This finding is supported by the established poor overall outcomes associated with acute myelitis, with only approximately one-third of patients experiencing a favourable outcome.^21^

A statistically significant relationship was noted between Asian ethnicity and peripheral neuropathy. The relationship between ethnicity and peripheral neuropathy in the context of COVID-19 has yet to be explored. However, peripheral neuropathy as a complication of diabetes has been found to be more prevalent among Caucasian patients^22^ and less common in those with Indo-Asian and African-Caribbean origins.^23^ Moreover, a statistically significant relationship was noted between Asian ethnicity and neuro-syndromic symptoms. Nonetheless, it is important to note that both of these relationships may have been influenced by the fact that the majority of the participants in the studies included were Asian and that a number of papers did not disclose the ethnicity of their participants.

Additionally, flu-like symptoms were statistically more prevalent in females, possibly because males have been found to have a higher risk of severe illness with COVID-19.^24^ Hyposmia and haemoptysis were statistically more prevalent in males. This is in contrast to several previous studies that found hyposmia to be more common in females with COVID-19 infection.^25–28^ However, our patient cohort was predominantly male (62%), which may have contributed to the differing results. Regarding haemoptysis, it is a very uncommon presentation that was only present in 10 patients.

ITU admission was found to have a statistically significant relationship with male sex, but not with age. A meta-analysis of patients with COVID-19 also demonstrated a relationship between sex and ITU admission, with male patients having almost three times the probability of requiring ITU admission compared to females.^29^ Surprisingly, our study did not determine any relationship between age and ITU admission. In contrast, another meta-analysis found that patients greater than 70 years old have a higher risk of needing intensive care.^30^ Furthermore, there was a statistically significant relationship between ITU admission and the symptoms of hyposmia/anosmia, headache, acute CVA, seizure, meningitis, and encephalopathies.

Treatment varied, with several different therapies and drug routes being used depending on the neurological manifestation and severity of the presentation. The most frequent treatments used were intravenous immunoglobulins (IVIG), followed by antibiotics such as azithromycin, antivirals, and hydroxychloroquine, with patients receiving treatment for a mean duration of 6 days. A systematic review assessing treatment strategies for COVID-19 similarly found antivirals, antimalarials, and antibiotics to be the mainstay of treatment.^31^ The frequency of IVIG can be attributed to its use in treating many different neurological conditions, most notably Guillain-Barre Syndrome, which was the fourth most common neurological complication reported in this review. Finally, it is important to consider that the COVID-19 pandemic is rapidly evolving and that treatment options are continually being trialled and developed.

Even though we established an abundance of studies for neurological symptoms, there appears to be a lack of studies regarding the psychiatric effects during and after COVID-19 infection. Nonetheless, all the studies we were able to identify reporting psychiatric effects have found depression and anxiety to be relevant, both during and after infection with COVID-19. In severe cases, there may even be a risk of patients attempting suicide. Compared to people who had flu or other respiratory tract infections, COVID-19 survivors were more likely to receive a diagnosis of anxiety of depression over the same period^17^. It was found that involving psychiatric care for these patients was effective in reducing their symptoms of anxiety and depression. Without proper psychiatric intervention, there is a risk that these psychiatric symptoms could increase the risk of suicidal ideation. Overall, it is recommended that psychiatric and/or psychological support should be available in hospitals to patients admitted to medical wards due to COVID-19, as well as in the community following recovery. This process may involve both the use of pharmacological and/or psychological interventions. Given the fact that COVID-19 survivors were at higher risk of receiving a diagnosis of dementia at 6-months follow-up, access to memory clinics should also be available to this group of patients. More studies examining the short-term and long-term psychiatric effects during and after COVID-19 infection are required in the future to obtain a better understanding of the symptoms, as well as to develop effective management strategies.

## Supporting information

Supplementary table 1 lists demographic information of the studies included in the review reporting neurological symptoms in the context of COVID-19

## Data Availability

All data referred to in the manuscript are publicly available.

https://www.crd.york.ac.uk/prospero/display_record.php?RecordID=203770

## SUPPLEMENTARY MATERIALS

Supplementary table 1 lists demographic information of the studies included in the review reporting neurological symptoms in the context of COVID-19 infection and/or recovery.

