## Supplementary table 1 lists demographic information of the studies included in the review reporting neurological symptoms in the context of COVID-19 for "Neurological, neuropsychiatric and psychiatric symptoms during COVID-19 infection and after recovery: a systematic review of observational studies"

**Supplementary Material 1: List of studies for neurological symptoms of COVID-19 included in the systematic review.**

| | Year published | Type of study | Country of publication | Sample size | % Female | % Male |
|---|---|---|---|---|---|---|
| Al Mazrouei et al | 2020 | Case Report | UAE | 1 | 0 | 100 |
| Al-Olama et al | 2020 | Case Report | UAE | 1 | 0 | 100 |
| AlKetbi, R. et al | 2020 | Case Report | UAE | 1 | 0 | 100 |
| Capelli, M. et al | 2020 | Case Series | Italy | 27 | 63 | 37 |
| Cotelli, M. et al | 2020 | Case Report | Italy | 1 | 100 | 0 |
| Daher, V. B. et al | 2020 | Case Report | Brazil | 1 | 0 | 100 |
| Dalakas, M. C. | 2020 | Case Series | .. | 11 | .. | .. |
| De Freitas F. et al | 2020 | Case Study | Brazil | 1 | 0 | 100 |
| Deliwala, S. et al | 2020 | Case Report | USA | 1 | 100 | 0 |
| Dugue, R. et al | 2020 | Case Report | .. | 1 | 0 | 100 |
| Espinosa, P. S. et al | 2020 | Case Report | .. | 1 | 0 | 100 |
| Fadakar, N. et al | 2020 | Case Report | Iran | 1 | 0 | 100 |
| Fan, S. et al | 2020 | Case Series | China | 86 | 32 | 63 |
| Farhadian, S. et al | 2020 | Case Report | USA | 1 | 100 | 0 |
| Farley, M. and Zuberi, J. | 2020 | Case Report | .. | 1 | 0 | 100 |
| Farzi, M. A. et al | 2020 | Case Report | Iran | 1 | 0 | 100 |
| Gilani, S. et al | 2020 | Case Series | Iran | 8 | 75 | 25 |
| Goldberg, M. F. et al | 2020 | Case Report | USA | 64 | 0 | 100 |
| Haddad, S. et al | 2020 | Case Report | USA | 41 | 0 | 100 |
| Haddadi, K. et al | 2020 | Case Report | Iran | 1 | 100 | 0 |
| Hamidi, A. et al | 2020 | Case Report | Iran | 1 | 0 | 100 |
| Hanafi, R. et al | 2020 | Case Report | .. | 1 | 0 | 100 |
| Hepburn, M. et al | 2020 | Case Series | USA | 2 | 0 | 100 |
| Hernández-Fernández, F. et al | 2020 | Case Series | Spain | 23 | 22 | 78 |
| Klironomos, S. et al | 2020 | Cohort Study | Sweden | 185 | 25 | 75 |
| Koleva, T. et al | 2020 | Retrospective Study | Bulgaria | 133 | .. | .. |
| Kremer, S. et al | 2020 | Retrospective Study | France | 64 | 33 | 67 |
| Lascano, A. M. et al | 2020 | Case Series | Switzerland | .. | 100 | 0 |
| Mahboob, S. et al | 2020 | Case Report | USA | 1 | 100 | 0 |
| Mao, L. et al | 2020 | Case Series | China | 214 | 59 | 41 |
| Najjar, S. et al | 2020 | Case Series | | 3 | 100 | 0 |
| Ottaviani, D. et al | 2020 | Case Report | Italy | 1 | 100 | 0 |
| Paniz-Mondolfi, A. et al | 2020 | Case Report | | 1 | 0 | 100 |
| Parsons, T. et al | 2020 | Case Report | USA | 1 | 100 | 0 |
| Paybast, S. et al | 2020 | Case Report | Iran | 2 | 50 | 50 |
| Pinna, P. et al | 2020 | Case Series | USA | 50 | 42 | 58 |
| Poncet-Megemont, L. et al | 2020 | Cohort Study | France | 139 | 33 | 67 |
| Reichard, R. R.et al | 2020 | Case Report | USA | 1 | 0 | 100 |
| Rogg, J. et al | 2020 | Case Report | | 1 | 1 | 100 |
| Romero Cantero, V. et al | 2020 | Case Series | | 2 | 0 | 100 |

| | | | | | | |
|---|---|---|---|---|---|---|
| Romero-Sánchez, C. et al | 2020 | Observational Study | Spain | 841 | 44 | 56 |
| Collantes, M. E. V. | 2020 | Systematic Review | Philippines | 6,335 | ·· | ·· |
| Chang, D. et al. | 2020 | | China | 13 | 23 | 77 |
| Chen, T. et al. | 2020 | Retrospective Study | China | 203 | 47 | 53 |
| Chung, M. et al. | 2020 | Retrospective study | China | 21 | 38 | 62 |
| Du, RH et al. | 2020 | Prospective study | China | 179 | ·· | ·· |
| Gupta, N. et a. | 2020 | Retrospective study | India | 21 | ·· | ·· |
| Kim et al. | 2020 | Cohort study | Korea | 28 | 46 | 54 |
| Lei, Z. et al. | 2020 | Cross sectional study | China | 20 | 50 | 50 |
| Li, X et al. | 2020 | Retrospective study | China | 548 | 49 | 51 |
| Liu, Y. et al. | 2020 | Retrospective | China | 12 | 33 | 67 |
| Pan, F. et al. | 2020 | Retrospective | China | 21 | 71 | 29 |
| Shen, L. et al. | 2020 | Retrospective | China | 119 | 53 | 47 |
| Shi, H. et al. | 2020 | Retrospective study | China | 81 | 48 | 52 |
| Song, F. et al. | 2020 | Retrospective study | China | 51 | 51 | 49 |
| Vu, D. et al. | 2020 | Case report | | 1 | 0 | 100 |
| Wang, R. et al. | 2020 | Retrospective study | China | 125 | 43 | 57 |
| Wang Z. et al. | 2020 | Retrospective study | China | 2 | 50 | 50 |
| Pongpirul, WA. et al. | 2020 | | Thailand | 11 | 45 | 55 |
| Wu, J. et al. | 2020 | Retrospective study | China | 80 | 47 | 53 |
| Xu, X. et al. | 2020 | Case series | China | 90 | 57 | 43 |
| Yang, X. et al. | 2020 | Retrospective study | China | 52 | 33 | 67 |
| Zheng, F. et al. | 2020 | Retrospective study | China | 161 | 50 | 50 |
| Correia, A. O. | 2020 | Systematic Review | Brazil | 409 | ·· | ·· |
| Moriguchi et al. | 2020 | Case Report | Japan | 1 | ·· | 100 |
| Poyiadji et al. | 2020 | Case Report | USA | 1 | 100 | |
| De Sanctis, P. | 2020 | Systematic Review | Italy | 18 | 44 | 56 |
| Toscano, G. et al | 2020 | Case series | Italy | 5 | 20 | 80 |
| Virani et al. | 2020 | Case report | USA | 1 | 0 | 100 |
| Zhao H. et al. | 2020 | Case report | China | 1 | 100 | 0 |
| Sedaghat and Karimi | 2020 | Case report | Iran | 1 | 0 | 100 |
| Alberti et al. | 2020 | Case report | Italy | 1 | 100 | 0 |
| Camdessanche et al. | 2020 | Case report | France | 1 | 0 | 100 |
| Padroni et al. | 2020 | Case report | Italy | 1 | 100 | 0 |
| El Otmani et al. | 2020 | Case report | Morocco | 1 | 100 | 0 |
| Coen et al. | 2020 | Case report | Switzerland | 1 | 0 | 100 |
| Galan et al. | 2020 | Case report | Spain | 1 | 0 | 100 |
| Marta-Enguita et al. | 2020 | Case report | Spain | 1 | 100 | 0 |
| Scheidl et al. | 2020 | Case report | Germany | 1 | 100 | 0 |

| | | | | | | |
|---|---|---|---|---|---|---|
| Juliao Caamano et al. | 2020 | Case report | Spain | 1 | 0 | 100 |
| Di Carlo, D. T. | 2020 | Systematic Review | Italy | 12,157 | 49.4 | 50.6 |
| Beltran-Corbellini A et al. | 2020 | Case-control study | Spain | 79 | .. | .. |
| Chen et al. | 2020 | Descriptive study | China | 99 | 32 | 68 |
| Duanmu Y et al. | 2020 | Cross-sectional study | USA | 100 | 44 | 56 |
| Feng et al. | 2020 | Retrospective study | China | .. | .. | .. |
| Giacomelli A et al. | 2020 | Cross-sectional | Italy | 59 | 32 | 68 |
| Helms et al. | 2020 | | France | 58 | .. | .. |
| Jain R. et al. | 2020 | Retrospective study | USA | 454 | 39 | 61 |
| Lechien et al. | 2020 | | France-Italy | 417 | 63 | 37 |
| Li K et al. | 2020 | Retrospective study | China | 83 | 47 | 53 |
| Mahammedi et al. | 2020 | Retrospective study | Italy | 108 | 36 | 64 |
| Qin et al. | 2020 | Retrospective study | China | 1,875 | 50 | 50 |
| Radmanesh et al. | 2020 | Retrospective Case Series | USA | 242 | 38 | 62 |
| Tian et al. | 2020 | Retrospective study | China | 262 | 52 | 48 |
| Wan et al. | 2020 | Case series | China | 135 | 47 | 53 |
| Wang et al | 2020 | Case series | China | 138 | 46 | 54 |
| Xu et al. | 2020 | Retrospective case series | China | 62 | 44 | 56 |
| Dinakaran, D. | 2020 | Systematic Review | | 399 | .. | .. |
| Yin et al. | 2020 | Case report | China | 1 | 0 | 100 |
| Ye et al. | 2020 | Case report | China | 1 | 0 | 100 |
| Alkeridy et al. | 2020 | Case report | Saudi Arabia | 1 | 0 | 100 |
| Duong et al. | 2020 | Case report | | 1 | 100 | 0 |
| Avula et al. | 2020 | Case series | | 4 | 75 | 25 |
| Ellul, M. A. et al | 2020 | Systematic Review | UK | .. | .. | .. |
| Bernard-Valnet et al | 2020 | Case series | Switzerland | 2 | 100 | 0 |
| Pilotto et al | 2020 | Case report | Italy | 1 | 0 | 100 |
| Sohal et al | 2020 | Case report | USA | 1 | 0 | 100 |
| Vollono et al | 2020 | Case report | Italy | 1 | 100 | 0 |
| Wong et al | 2020 | Case report | UK | 1 | 0 | 100 |
| Benussi et al | 2020 | Case series | Italy | 15 | | |
| Chacon-Aguilar et al | 2020 | Case report | Spain | 1 | 0 | 100 |
| Filatov A. et al | 2020 | Case report | USA | 1 | 0 | 100 |
| Garazzino S. et al | 2020 | Case series | Italy | 5 | .. | .. |
| Neerland et al | 2020 | Case report | Norway | 1 | 100 | 0 |
| Zhou et al | 2020 | Case report | China | 1 | .. | .. |
| Zhao K. et al | 2020 | Case report | China | 1 | 0 | 100 |
| Zanin et al | 2020 | Case report | Italy | 1 | 100 | 0 |
| Zhang T. et al | 2020 | Case report | USA | 1 | 100 | 0 |
| Abdelnour L. et al | 2020 | Case report | UK | 1 | 0 | 100 |
| Dinkin et al | 2020 | Case report | USA | 1 | 0 | 100 |

| | | | | | | |
|---|---|---|---|---|---|---|
| Gutierrez-Ortiz C. | 2020 | Case report | Spain | 1 | 0 | 100 |
| Pellitero S. | 2020 | Case report | Spain | 1 | 100 | 0 |
| Jin M | 2020 | Case report | China | 1 | 0 | 100 |
| Suwanwongse | 2020 | Case report | USA | 1 | 0 | 100 |
| Benezit | 2020 | Case series | France | 63 | ·· | ·· |
| Haehner | 2020 | Case series | Germany | 22 | ·· | ·· |
| Hornuss D. | 2020 | Case series | Germany | 18 | ·· | ·· |
| Klopfenstein | 2020 | Case series | France | 114 | ·· | ·· |
| Moien | 2020 | Case series | Iran | 59 | ·· | ·· |
| Vaira LA. | 2020 | Case series | | 72 | ·· | ·· |
| Al Saeigh | 2020 | Case report | USA | 2 | 50 | 50 |
| Beyrouti | 2020 | Case series | UK | 6 | 17 | 83 |
| Gonzalez-Pinto T. | 2020 | Case report | Spain | 1 | 100 | 0 |
| Klok FA | 2020 | Case series | Netherlands | 184 | 24 | 76 |
| Li, Y. et al. | 2020 | Retrospective study | China | 221 | 41 | 59 |
| Lodigiani | 2020 | Case series | Italy | 388 | 32 | 68 |
| Lushina, N. | 2020 | Case series | USA | 1 | 0 | 100 |
| Morassi, M | 2020 | Case series | Italy | 6 | 17 | 83 |
| Moshayedi P. | 2020 | Case report | USA | 1 | 0 | 100 |
| Oxley | 2020 | Case series | USA | 5 | 0 | 100 |
| Zhang Y. | 2020 | Case series | China | 3 | 33 | 67 |
| Zhai | 2020 | Case report | China | 1 | 0 | 100 |
| Sharifi-Razavi, A | 2020 | Case report | Iran | 1 | 0 | 100 |
| Garg, Ravindra Kumar. | 2020 | Systematic Review | ·· | 56 | ·· | ·· |
| Huang et al. | 2020 | Case Report | USA | 1 | 100 | 0 |
| Benameur et al. | 2020 | Case series | USA | 3 | 33 | 67 |
| McAbee et al. | 2020 | Case report | USA | 1 | 0 | 100 |
| Chaumont et al. | 2020 | Case report | Guadeloupe | 1 | 0 | 100 |
| Hayashi M et al. | 2020 | Case report | Japan | 1 | 0 | 100 |
| Afshar et al. | 2020 | Case report | Iran | 1 | 100 | 0 |
| Dixon et al. | 2020 | Case report | UK | 1 | 100 | 0 |
| Elkady et al. | 2020 | Case report | Egypt | 1 | 100 | 0 |
| Dogan et al. | 2020 | Case series | Turkey | 6 | 17 | 83 |
| Brun et al. | 2020 | Case report | France | 1 | 100 | 0 |
| Byrnes et al. | 2020 | Case report | USA | 1 | 0 | 100 |
| Novi et al. | 2020 | Case report | Italy | 1 | 100 | 0 |
| Nepal P. et al. | 2020 | Case report | USA | 1 | 0 | 100 |
| Balestrino R. et al | 2020 | Case report | Italy | 1 | 0 | 100 |
| Zayet S. et al. | 2020 | Case series | France | 2 | 0 | 100 |
| Somani et al. | 2020 | Case series | USA | 1 | 100 | 0 |
| Balloy et al. | 2020 | Case report | France | 1 | 0 | 100 |
| Anzalone et al. | 2020 | Case series | Italy | 4 | 50 | 50 |
| Sachs JR et al. | 2020 | Case study | USA | 1 | 0 | 100 |
| Kaya et al. | 2020 | Case report | Turkey | 1 | 0 | 100 |
| Franceschi et al. | 2020 | Case series | USA | 2 | 50 | 50 |
| Kishfy et al. | 2020 | Case series | USA | 2 | 50 | 50 |

| | | | | | | |
|---|---|---|---|---|---|---|
| Packwood et al. | 2020 | Case report | USA | 1 | 0 | 100 |
| Matos et al. | 2020 | Case report | Portugal | 1 | 0 | 100 |
| Ghannam, M. et al | 2020 | Systematic Review | USA | 82 | 18 | 82 |
| Wei H. et al | 2020 | Case study | China | 1 | 0 | 100 |
| Zhou B. et al. | 2020 | Case report | China | 1 | 100 | 0 |
| Valderrama EV. Et al. | 2020 | Case report | | 1 | 0 | 100 |
| Viguier A. et al. | 2020 | Case report | | 1 | 0 | 100 |
| Hughes et al. | 2020 | Case report | | 1 | 0 | 100 |
| Gupta, I. | 2020 | Systematic Review | | 1034 | 55 | 45 |
| Gane et al. | 2020 | Case report | UK | 1 | 0 | 100 |
| Eliezer et al. | 2020 | Case report | France | 1 | 100 | 0 |
| Spinato et al. | 2020 | | Italy | 202 | 52 | 48 |
| Munhoz, R. P. | 2020 | Systematic Review | | .. | .. | .. |
| Huang, C. et al. | 2020 | | China | 41 | 29 | 71 |
| Lu, L. et al. | 2020 | Retrospective study | China | 304 | 40 | 60 |
| Neishaboori, A. M. | 2020 | Systematic Review | | 1,643 | 41 | 59 |
| Chen, T. et al. | 2020 | Retrospective study | China | 274 | 38 | 62 |
| Nepal, G. | 2020 | Systematic Review | | .. | .. | .. |
| Karimi, N. et al. | 2020 | Case report | Iran | 1 | 100 | 0 |
| Qiu, H. et al. | 2020 | Cohort study | China | 36 | .. | .. |
| Zhou, F. et al. | 2020 | Cohort study | China | 191 | .. | .. |
| Guan, WJ. et al. | 2020 | | China | 1,099 | 49 | 51 |
| Fu, B. et al. | 2020 | Case series | China | 2 | 0 | 100 |
| Wan, Y. et al. | 2020 | Case report | China | 1 | 100 | 0 |
| Yan, CH. et al. | 2020 | Retrospective study | USA | 169 | .. | .. |
| Payus, A. O. | 2020 | Systematic Review | | .. | .. | .. |
| Xinhua et al. | 2020 | Case report | China | 1 | 0 | 100 |
| Pinzon, R. T. | 2020 | Systematic Review | Indonesia | 7,559 | .. | .. |
| Qian, GQ. et al. | 2020 | Case series | China | 88 | .. | .. |
| Hu et al. | 2020 | Case series | China | 24 | .. | .. |
| Wu, J. et al. | 2020 | Cross-sectional | China | 80 | .. | .. |
| Liu, K. et al. | 2020 | Cross-sectional | China | 137 | .. | .. |
| Guan, WJ. Et al. | 2020 | Cohort study | China | 1,590 | .. | .. |
| Zhang, X. et al. | 2020 | Cohort study | China | 645 | .. | .. |
| Han Y, et al. | 2020 | Cohort study | China | 25 | .. | .. |
| Wang, L. et al. | 2020 | Cohort study | China | 339 | .. | .. |
| Cai, Q. et al. | 2020 | Cohort study | China | 298 | .. | .. |
| Cao, J. et al. | 2020 | Cohort study | China | 102 | .. | .. |
| Yang, W. et al. | 2020 | Cohort study | China | 149 | .. | .. |
| Korea Centers for Disease Control and Prevention | 2020 | Case series | South Korea | 28 | .. | .. |
| Zhao, W. et al. | 2020 | Cohort study | China | 101 | .. | .. |
| Xu, YH. et al. | 2020 | Cohort study | | 50 | .. | .. |

| | | | | | | |
|---|---|---|---|---|---|---|
| Li, X. et al. | 2020 | Cohort study | China | 131 | ·· | ·· |
| Wang, X. et al. | 2020 | Cohort study | China | 1,012 | ·· | ·· |
| Rogers, J. P. | 2020 | Systematic Review | UK | 3,559 | ·· | ·· |
| Chen, G. et al. | 2020 | Cohort study | China | 21 | 19 | 81 |
| Zhang, B. et al. | 2020 | Cohort study | China | 82 | 38 | 62 |
| Qi, D. et al. | 2020 | Retrospective study | China | 267 | 44 | 56 |
| Huang, Y. et al. | 2020 | Cohort study | China | 36 | 31 | 69 |
| Leung et al. | 2020 | | China | 50 | 54 | 46 |
| Seyed Hamidreza Bagheri et al | 2020 | Cross-sectional study | Iran | 10,069 | 81.68 | 18.32 |
| Á. Beltrán-Corbellini et al | 2020 | Case control study | UK | 79 | 39.2 | 60.8 |
| R. Bernard-Valnet et al | 2020 | Case report | UK | 2 | 100% | 0 |
| Tao Chen et al | 2020 | Retrospective case series | UK | 274 | 38 | 62 |
| Lisa Duong et al | 2020 | Case report | USA | 1 | 100 | 0 |
| Asia Filatov et al | 2020 | Case report | USA | 1 | 0 | 100 |
| Consuelo Gutiérrez-Ortiz et al | 2020 | Case series | Germany | 2 | 0 | 100 |
| Julie Helms et al | 2020 | Case series | USA | 58 | ·· | ·· |
| Jerome R Lechien et al | 2020 | Case series | Germany | 417 | 63.1 | 36.9 |
| Yanan Li et al | 2020 | Case series | UK | 219 | 59.4 | 40.6 |
| Lu Lu et al | 2020 | Case series | USA | 304 | 40.1 | 59.9 |
| Ling Mao et al | 2020 | Case series | USA | 214 | 59.3 | 40.7 |
| Shima T Moein et al | 2020 | Case-control study | USA | 60 | 33.3 | 66.6 |
| Takeshi Moriguchi Takeda et al | 2020 | Case report | Netherlands | 1 | 0 | 100 |
| Marina Padroni et al | 2020 | Case report | Germany | 1 | 100 | 0 |
| Neo Poyiadji et al | 2020 | Case report | USA | 1 | 100 | 0 |
| Z. Sedaghata and N. Karimib | 2020 | Case report | USA | 1 | 0 | 100 |
| A. Sharifi-Razavi et al | 2020 | Case report | Netherlands | 1 | 0 | 100 |
| Gianpaolo Toscano et al | 2020 | Case series | USA | 5 | 20 | 80 |
| Catello Vollono et al | 2020 | Case report | UK | 1 | 100 | 0 |
| Carol H. Yan et al | 2020 | Cross-sectional | USA | 59 | 49.2 | 49.2 |
| Mingxiang Ye et al | 2020 | Case report | USA | 1 | 0 | 100 |
| Rong Yin et al | 2020 | Case report | USA | 1 | 0 | 100 |
| Tianshu Zhang et al | 2020 | Case report | USA | 1 | 100 | 0 |
| Hua Zhao et al | 2020 | Case report | UK | 1 | 100 | 0 |
| Kang Zhao et al | 2020 | Case report | ·· | 1 | 0 | 100 |
| Giacomo Spinato et al | 2020 | Case series | USA | 202 | 51 | 49 |
| François Bénézit et al | 2020 | Case series | UK | 259 | ·· | ·· |
| Sancho-Saldaña et al | 2020 | Case Report | UK | 1 | 100 | 0 |
